# Seroprevalence of Dengue, Chikungunya and Zika at the epicenter of the congenital microcephaly epidemic in Northeast Brazil: a population-based survey

**DOI:** 10.1101/2023.03.27.23287774

**Authors:** Cynthia Braga, Celina M.T. Martelli, Wayner V. Souza, Carlos F. Luna, Maria de Fatima P.M. Albuquerque, Carolline A. Mariz, Carlos A.A. Brito, Carlos Frederico C.A. Melo, Roberto D. Lins, Jan Felix Drexler, Thomas Jaenisch, Ernesto T.A. Marques, F.T. Viana Isabelle

**Affiliations:** Department of Parasitology, Aggeu Magalhães Institute, Oswaldo Cruz Foundation, Recife, Pernambuco, Brazil; Department of Public Health, Aggeu Magalhães Institute, Oswaldo Cruz Foundation, Recife, Pernambuco, Brazil; Department of Virology and Experimental Therapeutics, Aggeu Magalhães Institute, Oswaldo Cruz Foundation, Recife, Pernambuco, Brazil; Department of Clinical Medicine, Federal University of Pernambuco, Recife, Pernambuco, Brazil; Pan American Health Organization, Brasília, Federal District, Brazil; Charité—Universitätsmedizin Berlin, Corporate Member of Freie Universität Berlin and Humboldt-Universität zu Berlin, Institute of Virology, Berlin, Germany; German Centre for Infection Research (DZIF), associated partner site Charité, Berlin, Germany; Section Clinical Tropical Medicine, Department of Infectious Diseases, Heidelberg University Hospital, Germany; German Centre for Infection Research (DZIF), Heidelberg Site, Heidelberg, Germany; Center for Global Health, Colorado School of Public Health, Aurora, Colorado, USA; Department of Infectious Diseases and Microbiology, University of Pittsburgh, Pittsburgh, PA, USA

**Keywords:** Zika Infection, Chikungunya Virus Infection, Dengue, Seroepidemiological Study, Risk Factor, Brazil, ZIKV, CHIKV, DENV, Seroprevalence.

## Abstract

**Background:** The Dengue viruses (DENV) serotypes 1, 2, 3 and 4 were re-introduced in the Northeast Brazil from the 1980’s until 2010’s. Zika (ZIKV) and Chikungunya (CHIKV) viruses were introduced around 2014 and caused large outbreaks in 2015 and 2016. However, the true extent of the ZIKV and CHIKV outbreaks and the risk factors associated with exposure remain vague.

**Methods:** We conducted a stratified multistage household serosurvey among residents aged between 5 and 65 years in the city of Recife, Northeastern Brazil, from August 2018 to February 2019. The city neighborhoods were stratified according to high, intermediate, and low socioeconomic strata (SES). Previous ZIKV, DENV and CHIKV infections were detected by IgG based enzyme linked immunosorbent assays (ELISA) . Recent ZIKV and CHIKV infections were assessed through IgG3 and IgM ELISA, respectively. Design-adjusted seroprevalence were estimated by age group, sex, and SES. The ZIKV seroprevalence was adjusted to account for the cross-reactivity with dengue. Individual and household-related risk factors were analyzed through regression models to calculate the force of infection. Odds Ratio (OR) were estimated as measure of effect.

**Principal findings:** A total of 2,070 residents were investigated. The forces of infection for high SES were lower for all three viruses as compared to low SES. Overall, DENV seroprevalence was 88.7% (CI95%:87.0-90.4), (81.2% (CI95%:76.9-85.6) in the high SES and 90.7% (CI95%:88.3-93.2) in the low). The overall adjusted ZIKV seroprevalence was 34.6% (CI95%:20.0-50.9), (47.4% (CI95%:31.8-61.5) in the low SES and 23.4% (CI95%:12.2-33.8) in the high). CHIKV seroprevalence was 35.7% (CI95%:32.6-38.9), (38.6% (CI95%:33.6-43.6) in the low SES and 22.3% (CI95%:15.8-28.8) in the high). ZIKV seroprevalence increased with age while CHIKV seroprevalence was almost constant through age. The serological markers of recent infections for ZIKV and CHIKV were 1.5% (CI95%:0.1-3.7) and 3.5% (CI95%:2.7-4.2) respectively.

**Conclusions:** Our results confirmed continued DENV transmission and intense ZIKV and CHIKV transmission during the 2015/2016 epidemics followed by ongoing low-level transmission. The study also highlights that a significant proportion of the population is likely still susceptible to be infected by ZIKV and CHIKV, raising questions on herd immunity and antibody detection thresholds and the reasons underlying cease of the ZIKV epidemic in 2017/18.

**Author summary:** The extent and population burden of the Zika and Chikungunya epidemics in Northeast Brazil remains speculative since seroprevalence studies have often been restricted to specific populations and limited by ZIKV and DENV antibody cross-reactivity. Here we conducted a seroepidemiologicla study in the city of Recife, a metropolitan area in Northeastern Brazil using a design stratified by socio-economic status (SES). We also determined the sensitivity and specificity of the assays used and selected optimum cut-offs, which were and later confirmed by selecting a subset of samples to on which more specific virus neutralizations tests were performed. The result indicated that 89% of the population (older than 5 years of age) had previous dengue infection, compatible with our previous serosurvey. The assay sensitivity and specificity seroprevalences for ZIKV was 34.6% and CHIKV 35.7%, indicating high transmission during the outbreaks (2015/2016). Interestingly, the age distribution profiles of ZIKV and CHIKV seroprevalence were remarkably different. These differences cannot be explained by differences in mosquito exposure alone. Future research will need to be conducted to better explain the differences we found for the age distributions.

## Introduction

Arthropod-borne diseases, especially dengue, chikungunya and Zika, have presented a major public health problem in the Americas [1]. Brazil has been responsible for more than 90% of the reported cases of these arboviruses in this region in the last decades [2].

The recent introduction of Zika virus (ZIKV, genus *Flavivirus*, family *Flaviviridae*) in the Northeast Region of Brazil, between 2013 and 2015, was followed by its rapid spread to other regions causing significant increase in the number of cases of Guillain-Barré syndrome in adults and congenital microcephaly secondary to maternal ZIKV infection during pregnancy [3, 4]. The Zika outbreak was followed by an outbreak of the chikungunya virus (CHIKV, genus *Alphavirus*, family *Togaviridae*) in 2016 [5]. Data from the Brazilian Ministry of Health show that the states of Bahia, Ceara and Pernambuco, all in the Northeast region, were the most affected in Brazil [6, 7].

Population-based seroprevalence surveys stratified by age and geographic areas have been considered one of the current research priorities, due to their ability to estimate variations in the level of exposure of the population according to time, age, and environment [8, 9]. These studies provide much anticipated granular data that will allow to infer more precisely the susceptibility and level of immunity in a given population [8].

In Brazil, the true magnitude of the ZIKV and CHIKV outbreaks and the extent of the ensuing low-level transmission is still not well known [10]. The city of Recife, a large urban center marked by profound social inequalities in the northeast region of Brazil, has been affected by successive arbovirus epidemics since the introduction of the dengue virus (DENV) in the 1980s [11]. A dengue serosurvey conducted between 2005 and 2006 in three socioeconomically distinct neighborhoods of Recife estimated an overall prevalence above 80% [12], and showed an inverse association between the force of infection and socioeconomic status (SES). Between 2015 and 2016, with the emergence of ZIKV and CHIKV in Brazil, more than 50,000 cases of dengue, Zika and chikungunya were registered in this city [13, 14], which also accounted for around 90% of the cases of microcephaly attributed to ZIKV in Brazil [15, 16], of which97% occurred in babies of mothers with low SES. The reason for this disproportional distribution remains unclear, and one hypothesis is that pregnant women of high SES were much less exposed to Zika. We now carried out a detailed population-based survey in the whole city to determine the levels of exposure to ZIKV, DENV and CHIK by age and SES using validated serological tests.

## Methods

### Ethical Statement

The research project was reviewed and approved by the Research Ethics Committee of the Aggeu Magalhães Institute (Fiocruz, Pernambuco) (CAEE: 79605717.9.0000.5190, report number 2.734.481). Data collection was conducted after the participants or their legal guardians (if under 18 years old) were informed about the objectives of the study, read, and sign the consent form. Participants aged 5 to 18 years provided oral and/or written assent. All participants had access to the results of the laboratory tests. Personal information was removed prior to data analysis.

### Study design, population, and settings

The seroprevalence survey was conducted using a stratified multistage cluster sampling design involving residents aged between 5 and 65 years old, from August 2018 to February 2019. Recife, the capital city of Pernambuco state, has a territorial area of 218.8 km² (divided in 94 neighborhoods), and an estimated population of approximately 1.6 million inhabitants and a demographic density of 7,037.6 inhabitants/km². The city is classified as the 12th most densely populated urban area in Brazil. Around 40% of its population lives in poverty, with a monthly income of up to ½ minimum wage and 30% of households are in areas with inadequate sanitation [17].

### Sampling

The city’s neighborhoods were stratified in three economic strata based on the information of the family’s head income per census tract obtained in 2010 Demographic Census. The census tract (CT) is the smallest territorial unit for obtaining population data and has approximately 300 households or approximately 1,000 inhabitants [18].

Briefly, the division of the city’s territory at neighborhood level was performed by initially calculating the percentage of households with family’s heads without income or with monthly income <2 minimum wage (MW) per CT. Subsequently, these CT were aggregated at neighborhood level which were classified into four clusters relatively homogeneous with respect to socioeconomic status, using the k-means clustering technique and ANOVA test [19]. Further details on the methodology for stratifying the city’s territory were described elsewhere [20]. In this study, the high socioeconomic stratum was formed by merging the two clusters of neighborhoods with the highest household income (Clusters 1 and 2), due to the small population size in both clusters. The 3rd and 4th clusters were classified as intermediate and low socioeconomic strata, respectively.

The population sample size in each stratum was calculated considering an expected seroprevalence of 30% in the high socioeconomic strata and of 40% in the intermediate and low socioeconomic strata, respectively; absolute error of 4%; design effect = 1.5 and, 95% confidence level, yielding a sample of 2,500 participants: 760 residents in the high socioeconomic stratum, and 870 in both the intermediate and low strata.

The number of CT (primary sampling units) to be selected in each stratum was determined considering an estimated prevalence of arbovirus infected individuals (60%, in the high stratum; and 80%, in both intermediate and low strata) per CT, yielding a sample of 40 CT in the high socioeconomic stratum and 30 CT in the intermediate and low strata, respectively, which represents a total of 100 CT across the city.

The number of households to be selected in each CT was determined based on the estimate of 3.5 inhabitants per household eligible for the study, obtaining a sample of 716 households: 218 households, in the high socioeconomic stratum, and 249, in the intermediate and low strata. The selection of the participants was conducted in two stages. First, we randomly selected the CT (sampling units) in the strata (first stage) and next (second stage), the households (including all residents in the study age group) within the CT using the free R [21] package Amostra Brasil for household sampling (with their respective geographic coordinates) in Brazilian municipalities from the IBGE database [22]. Fig 1 shows the spatial distribution of the household sampled by socioeconomic strata in the city.

**Figure 1.**
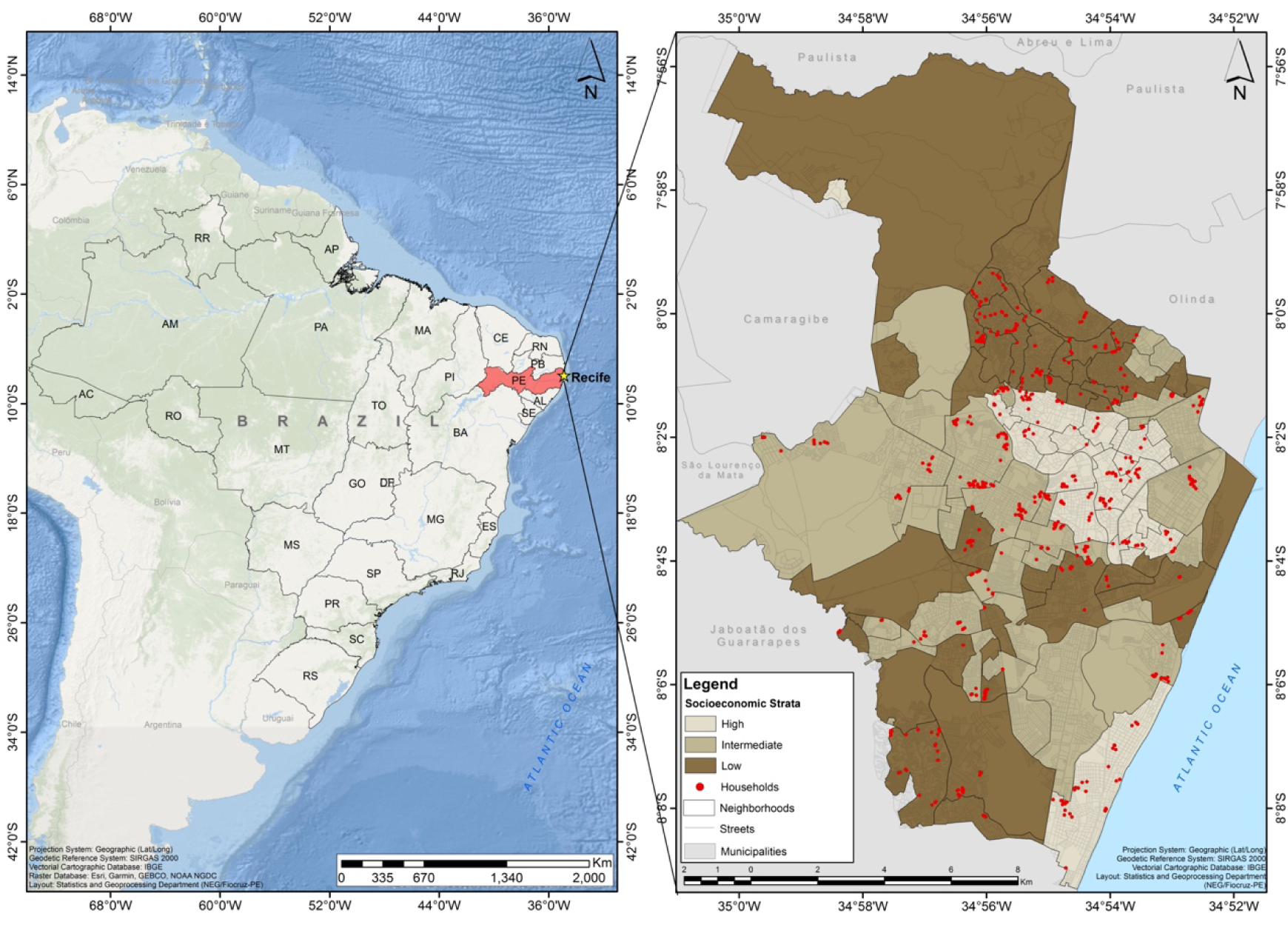
Spatial distribution of the selected households according to socioeconomic strata. Recife, Brazil.

### Data collection

The field team, comprised of interviewers and phlebotomists, was trained to ensure standardization and data quality. Home visits were initially performed to inform the head of the family about the objectives of the study and to invite the eligible residents to participate. Individual and household information were collected using standardized questionnaires (S1 Supplemental Material). After the interview, a venous blood samples of the participants (8 mL of adults and 5 mL of the children aged up to 9 years) was collected to perform the serological tests. Samples were collected in vacuum tubes with clot activator for 30 minutes at room temp and then placed in a container at 4°C. Urine and hair samples were also collected from the participants that showed signs of recent infection and transported to the Department of Virology at the Aggeu Magalhães Institute - FIOCRUZ, where they were processed and stored at -70°C until use.

### Laboratorial procedures

Anti-CHIKV IgG and IgM antibodies were detected through commercial ELISA kits (Euroimmun, Lubeck, Germany). The results of both tests were interpreted according to the manufacturer instructions (CHIKV IgG or IgM absorbance at 450 nm/calibrator ratios were considered negative at <0.8, indeterminate at ≥0.8 to <1.1, and positive at ≥1.1). All samples with indetermined results were retested using the same commercial kits, and the obtained results were considered as final.

The anti-ZIKV immune response was assessed by detecting IgG through a commercial ELISA kit (Euroimmun, Lubeck, Germany) [23]. Aiming to overcome the possible cross-reactivity between anti-DENV and anti-ZIKV immune responses, the cut-off of this test was redefined using a panel of 140 well-characterized serum samples from acute and convalescent samples of PCR confirmed DENV infection cases collected years prior to the Zika outbreak and samples from PCR confirmed cases of ZIKV infections. Results were interpreted as positive for ZIKV IgG when the sample absorbance at 450nm/calibrator ratio ≥1.35. Based on this cut-off point, the estimated sensitivity and specificity of this test were 86 % (95%CI: 72-95%) and 72% (95%CI: 65-79%), respectively (S2 – Supplemental Material).

Recent ZIKV infections were determined by detecting IgG3 against the ZIKV NS1 protein through an in-house ELISA [24–26]. Results were interpreted as positive for ZIKV IgG3 when the sample absorbance at 450nm/DENV recent infection control ratio ≥1.14. According to this cut-off, the estimated sensitivity and specificity of this test were 81% (95CI%: 60%-95%) and 93% (95%CI: 88%-96%), respectively (S2-Supplemental Material). The IgG3 ELISA detailed protocol is described in the Supporting Information (S2 Supplemental Material). The choice of this test over the commercially available ZIKV IgM kit (Euroimmun, Lubeck, Germany) was due to the lower sensitivity of the latter in our analysis using a well-characterized serum panel, where the lower sensitivity of this test in a scenario of flavivirus co-circulation has been documented [27, 28].

The accuracy of the ZIKV serology results was validated by performing a blind plaque reduction neutralization test (PRNT) of a subset of 156 randomly selected serum samples and compared with the other serological tests. The PRNT was performed following a modified protocol described in detail elsewhere [29] and neutralization was assessed against the ZIKV local strain (BR-PE243/2015). The cut-off for PRNT positivity was defined based on a 50% reduction in plaque counts (PRNT50), and ZIKV-specific antibody titers were estimated using a four-parameter non-linear regression. Samples were considered positive when the PRNT50s were ≥1:100 for ZIKV (S2 Supplemental Material) corroborating sensitivity and specificity data. The duration of Zika binding and neutralization antibodies two years after infection have been also evaluated in a previous study that showed significant decay of antibody levels but very few seroreversions [30].

Previous exposure to DENV was assessed by detecting IgG against the DENV 1-4 NS1 proteins through an in-house indirect ELISA, as described elsewhere [31]. Samples were considered positive for DENV 1-4 IgG when sample absorbance at 450nm/positive control ratio ≥3.62, corresponding to a sensitivity of 96% (95% CI:90-99%) and specificity of 71% (95% CI: 58-82%) (S2 Supplemental Material).

### Laboratorial classification of samples

Serological and/or neutralization positive samples for ZIKV or only for CHIKV were classified as ZIKV+ and CHIKV+, respectively. Negative samples for ZIKV and CHIKV tests and positive for DENV 1-4 ELISA test were classified as ZIKV-/CHIKV-/DENV+. Samples that were negative for all arbovirus tested were classified as ZIKV-/CHIKV-/DENV-.

### Exposure variables

We considered exposure variables on household level as well as on individual level. Household level: residents per bedroom, type of household, sewerage destination, water supply, frequency of water supply, garbage collection, sociodemographic characteristics of the head of the family. Individual level: age (in years), age group (5-14, 15-24, 25-34, 35-44, 45-54, 55-65), gender, self-reported skin color, schooling (≥13 years old), previous dengue infection, use of repellent, previous dengue exposure, vaccination for Yellow fever virus or DENV. Participants who reported fever or skin rash in the 30 days prior to the interview were asked about other clinical manifestations suggestive of arbovirus infection (dengue, chikungunya or Zika) and collected urine and hair samples (S1 Supplementary Material and Table 1).

**Table 1:** Characteristics of the selected households according to socioeconomic strata of the city of Recife, 2018-2019.

| Characteristics | Total | Socioeconomic Strata |  |  | p-value |
| --- | --- | --- | --- | --- | --- |
|  |  | High | Intermediate | Low |  |
| **Studied households. n (%)** | 892 (100.0) | 231 (25.9) | 349 (39.1) | 312 (35.0) |  |
| **Residents per bedroom, mean (se)** | 1.61 (0.03) | 1.36 (0.04) | 1.65 (0.04) | 1.75 (0.04) | <0.001 |
| **Type of household, n (%)** |  |  |  |  |  |
| Apartment | 202 (22.8) | 146 (63.5) | 67 (19.4) | 12 (3.8) | <0.001 |
| House | 682 (77.2) | 84 (36.5) | 279 (80.6) | 300 (96.2) |  |
| **Sewerage, n (%)\*** |  |  |  |  |  |
| Public network | 474 (56.0) | 157 (75.9) | 204 (60.7) | 116 (37.7) |  |
| Other waste destinations | 373 (44.0) | 49 (24.1) | 132 (39.3) | 192 (62.3) | <0.001 |
| **Water supply, n (%)** |  |  |  |  |  |
| Public network | 755 (85.0) | 157 (68.3) | 319 (92.0) | 379 (89.4) |  |
| Other sources (well, others) | 133 (15.0) | 73 (31.7) | 27 (7.8) | 33 (10.6) | <0.001 |
| **Regular water supply, n (%)\*\*** | 667 (75.2) | 157 (89.1) | 281 (81.2) | 181 (58.2) | <0.001 |
| **Household garbage collection. n (%)** | 832 (93.7) | 227 (98.7) | 316 (91.3) | 289 (92.6) | <0.001 |
| **Characteristics of the head of the family, n (%)** |  |  |  |  |  |
| **Sex** |  |  |  |  |  |
| Female | 484 (54.7) | 98 (49.4) | 178 (57.3) | 208 (55.4) |  |
| Male | 401 (45.3) | 101 (50.6) | 133 (42.7) | 167 (44.6) | 0.298 |
| **Self-reported race/skin color** |  |  |  |  |  |
| Mixed race (Brown) | 451 (50.6) | 92 (39.8) | 164 (47.0) | 195 (62.5) |  |
| Black | 125 (14.0) | 20 (8.7) | 50 (14.3) | 55 (17.6) |  |
| White | 283 (31.7) | 107 (46.3) | 122 (35.0) | 54 (17.3) |  |
| Others/not informed/ignored | 33 (3.7) | 12 (5.2) | 13 (3.7) | 8 (2.6) | 0.001 |
| **Monthly income in minimum wages** |  |  |  |  |  |
| ≤ 2 | 550 (62.4) | 57 (28.7) | 181 (58.7) | 312 (85.3) |  |
| 2-4 | 185 (21.0) | 45 (22.6) | 84 (27.2) | 56 (15.1) |  |
| 4-20 | 146 (16.6) | 97 (48.7) | 44 (14.2) | 6 (1.6) | 0.001 |
| **Schooling** |  |  |  |  |  |
| University | 238 (27.0) | 133 (67.2) | 86 (27.6) | 19 (5.1) |  |
| High school | 299 (33.9) | 36 (18.3) | 112 (35.9) | 151 (40.5) |  |
| Fundamental | 345 (39.1) | 28 (14.4) | 113 (36.5) | 203 (54.3) | 0.001 |

### Statistical analysis

Double data entry and data consistency analysis were performed using REDCap electronic data capture tools [32] hosted at Heidelberg University, Germany. The analyzes were performed using the software R version 4.0.27 [21].

### Seroprevalence estimates

The seroprevalence of dengue (IgG), Zika (IgG and/or IgG3) and chikungunya (IgG and/or IgM) and their respective 95% confidence intervals (95%CI) were estimated according to age group and sex for each socioeconomic stratum (high, intermediate, and low). These estimates were weighted by the effect of the sample design using the “survey” package, version 4.1-1 (http://r-survey.r-forge.r-project.org/survey/) (S3 Supplementary Material). Considering the low accuracy of the Zika serological tests (due to the cross reactivity with anti-DENV antibodies) in areas of cocirculation of DENV and ZIKV, we estimated the prevalence of anti-ZIKV IgG and IgG3 through a Bayesian method for estimation of true prevalence from apparent prevalence obtained by testing individual samples [33, 34]. The library prevalence (which provides frequentist and Bayesian methods useful in prevalence assessment studies) was used assuming a uniform range of sensitivity and specificity between using a range corresponding to the 95% CI of the sensitivities and specificities calculated for the Zika IgG and IgG3 tests. The beta distribution was defined based on a priori distribution of the true prevalence. According to these estimates, 10,000 first iterations were discarded and the average of the remaining 20,000, together with their respective standard deviations, were used to estimate the true prevalence of Zika by socioeconomic stratum, sex, and age group. The analysis of the convergence of estimates was performed through the Multivariate BGR statistic proposed by Gelman and Rubin [35] and improved by Brooks and Gelman [36]. We used the Pearson’s Chi-square test with Rao-Scott correction [37] to compare the prevalence of Dengue, Zika and Chikungunya within and between socioeconomic strata and the *t*-Student or ANOVA tests to compare the means of the true prevalence of Zika estimated by the Bayesian method. This method proposes an adjustment of the seroprevalence estimates taking the imprecision of the calculated sensitivity and specificity. In this case we used the 95% CI of tests used

The force of dengue, Zika and chikungunya infection was estimated in each socioeconomic stratum assuming a constant risk of arbovirus exposure in the study population with permanent seroconversion. The model was fitted by the effect of the sample design from a generalized linear model (GLM) with serostatus as the outcome variable (anti-ZIKV IgG and/or IgG3 positive; anti-DENV IgG; anti-CHIKV IgM and/or IgG), a quasibinomial distribution family, complementary logit link function, and the age as exposure variable [38].

### Risk factor analysis for Zika and Chikungunya

The associations of the exposure variables with the seropositivity of Chikungunya and Zika were analyzed through hierarchical multiple regression analysis. Initially, univariate analysis was performed, calculating the crude odds ratio (OR) and their respective 95% CI for each block of variables (individual and household characteristics). The independent variables associated with the outcome at a significance level of p <0.25 were included in the multivariate logistic regression model for their respective block. The variables which remained that showed a statistically significant association with the outcome in the multiple regression models, within each block, were brought together in a new multivariate model to obtain the final model.

The selection method of variables used in each model regression was based in the Akaike Information Criterion (AIC).

## Results

### Characterization of the study population

A total of 2,691 residents of the visited households were eligible: 654 in the high; 1,037 in the intermediate; and 1,000 in the low SES stratum. Reasons for households not to participate included the difficulty in accessing the household (in general, an apartment) or(ii)refusals, which were more frequentin the high SES (52.4%) compared to the intermediate (18.8%) and low strata (13.6%). Among the eligible residents interviewed and consented for venous blood collection, 480 (73.3%) were from the high stratum; 815 (78.6%) from intermediate; and 775 (77.5%) from the low stratum (Fig 2)

**Figure 2.**
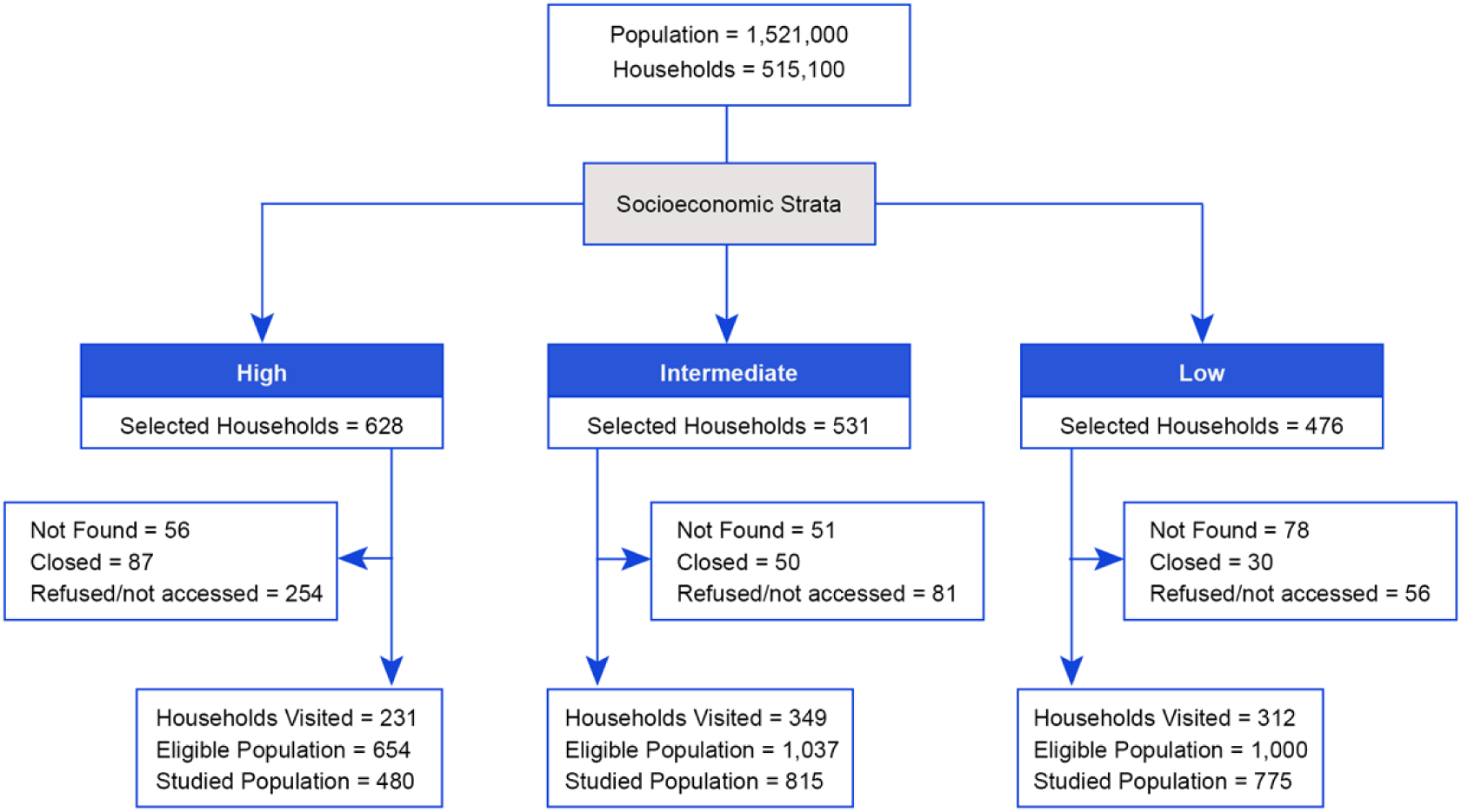
Population, census tracts and calculated samples by socioeconomic strata. Recife, Brazil, 2018-2019.

Table 1 shows the main characteristics of the studied households according to SES. We observed a gradient in the average number of residents per bedroom that increased from the high SES (1.61±0.03) to the low stratum (1.75±0.04). About 60% of the households in the high SES lived-in high-rise apartments, while most households in the low stratum were ground level houses. Almost half of the households had no access to public sewage, with a higher proportion among those in the low stratum (62.3%). More than half of the households had irregular water supply in the low stratum (41.8%). Near 30% of the households were supplied by artesian well in the high stratum. Heads of family from the high stratum had a high level of education and income compared to those from the intermediate and low strata.

### Seroprevalence of Dengue, Zika and Chikungunya

Of 2,070 participants, 1,837 had serological markers of previous DENV infection (anti-NS1 IgG-ELISA), that corresponded to an overall weighted prevalence of 88.7% (95%CI: 87.0%-90.4%), consistent with the seroprevalence levels determined in 2005/6 [12, 39]. Dengue seroprevalence ranged from 81.2% (95% CI: 76.9%-85.6%), in the high SES, to 90.7% (95%CI: 88.3%-93.2%), in the low SES (Fig 3, Table 2).

**Figure 3:**
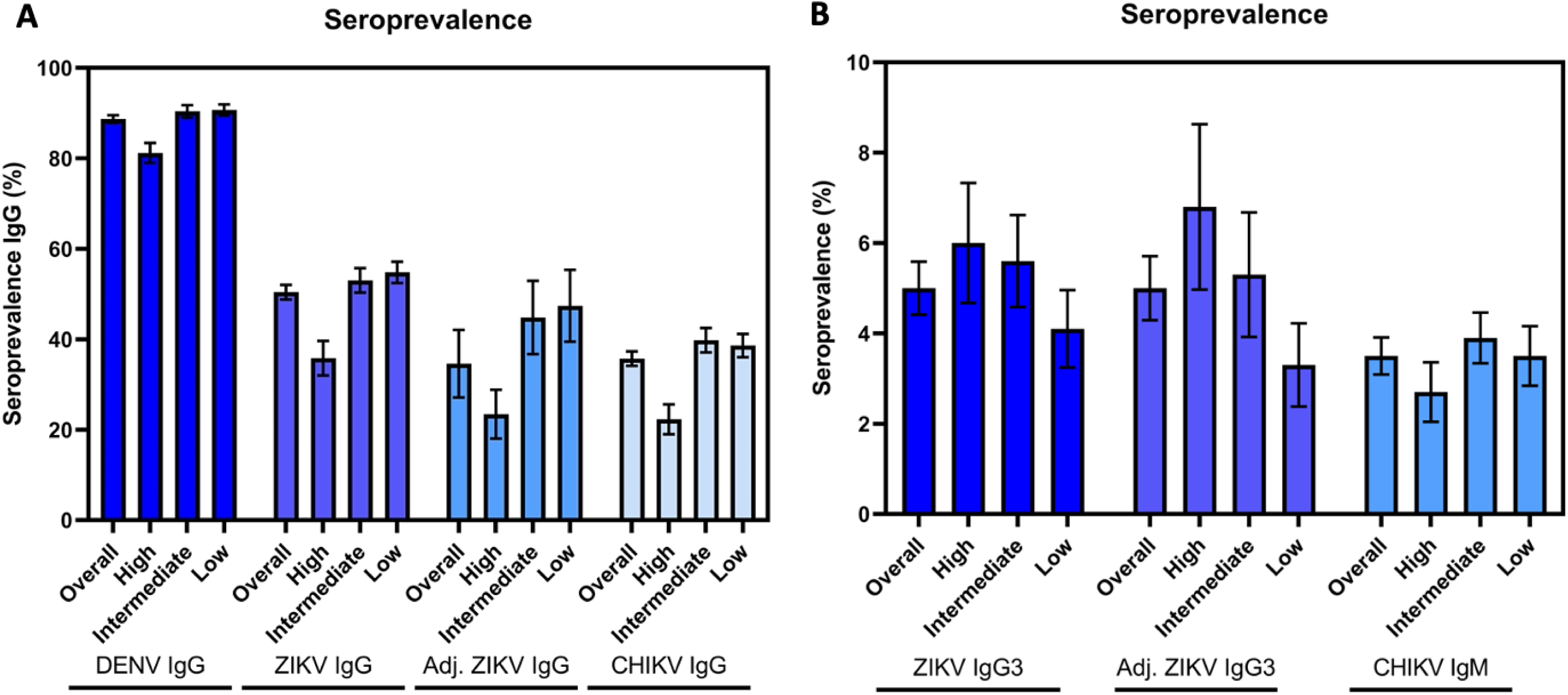
(A) Weighted IgG seroprevalence of Chikungunya Zika and Dengue including adjusted seroprevalence of ZIKA and (B) weighted seroprevalence of Chikungunya and Zika markers of recent infection (IgG3 and IgM) according to socioeconomic strata. Recife, Brazil, 2018-2019. Error bars indicate the standard deviations.

**Table 2.** –Weighted seroprevalence of Dengue (anti-DENV IgG-ELISA), Zika (Anti-ZIKV IgG e Anti-ZIKV IgG3) and Chikungunya (Anti-CHIKV Ig G e Anti-CHIKV Ig M) according to socioeconomic strata, sex and age group. Recife, Northeast of Brazil, 2018-2019.

|  | Examined | Dengue |  | Zika |  |  |  | Chikungunya |  |  |  |
| --- | --- | --- | --- | --- | --- | --- | --- | --- | --- | --- | --- |
|  |  | IgG-ELISA<sup>1</sup> (in house) |  | Anti-ZIKV IgG<sup>1</sup> |  | Anti-ZIKV IgG3<sup>1</sup> |  | Anti-CHIKV IgG<sup>1</sup> |  | Anti-CHIKV IgM<sup>1</sup> |  |
|  |  | Pos | Prev (CI95%) | Pos | Prev (CI95%) | Pos | Prev (CI95%) | Pos | Prev (CI95%) | Pos | Prev (CI95%) |
| **Overall** | 2,070 | 1,837 | 88.7 (87.0-90.4) | 1,043 | 50.4 (47.2-53.6) | 104 | 5.0 (3.9-6.2) | 740 | 35.7 (32.6-38.9) | 72 | 3.5 (2.7-4.2) |
| **Socioeconomic strata** |  |  |  |  |  |  |  |  |  |  |  |
| High | 416 | 338 | 81.2 (76.9-85.6) | 149 | 35.8 (28.3-43.3) | 25 | 6.0 (3.4-8.7) | 93 | 22.3 (15.8-28.8) | 11 | 2.7 (1.4-4.0) |
| Intermediate | 726 | 657 | 90.4 (87.7-93.2) | 385 | 53.0 (47.7-58.4) | 41 | 5.6 (3.6-7.7) | 289 | 39.8 (34.5-45.0) | 29 | 3.9 (2.8-5.1) |
| Low | 928 | 842 | 90.7 (88.3-93.2) | 509 | 54.8 (50.2-59.5) | 38 | 4.1 (2.4-5.8) | 358 | 38.6 (33.6-43.6) | 32 | 3.5 (2.2-4.8) |
| **Sex** |  |  |  |  |  |  |  |  |  |  |  |
| Female | 1,212 | 1,099 | 90.7 (88.6-92.7) | 616 | 50.8 (47.3-54.4) | 61 | 5.1 (3.7-6.4) | 439 | 36.2 (32.8-39.7) | 51 | 4.2 (3.0-5.4) |
| Male | 858 | 738 | 86.0 (83.1-88.8) | 427 | 49.7 (45.8-53.6) | 43 | 5.0 (3.3-6.7) | 301 | 35.0 (31.0-39.0) | 21 | 2.5 (1.5-3.5) |
| **Age Group (in years)** |  |  |  |  |  |  |  |  |  |  |  |
| 5-14 | 264 | 140 | 52.9 (45.8-60.1) | 65 | 24.6 (18.4-30.9) | 11 | 4.4 (1.7-7.0) | 80 | 30.5 (24.4-36.6) | 8 | 3.2 (1.1-5.2) |
| 15-24 | 357 | 319 | 89.3 (85.3-93.2) | 162 | 45.3 (38.8-51.9) | 14 | 4.0 (1.7-6.3) | 121 | 34.0 (28.7-39.3) | 16 | 4.5 (2.3-6.7) |
| 25-34 | 323 | 300 | 92.9 (90.1-95.8) | 160 | 49.7 (42.9-56.4) | 13 | 4.0 (1.7-6.3) | 118 | 36.6 (31.3-41.9) | 13 | 4.0 (2.2-5.9) |
| 35-44 | 375 | 360 | 96.0 (93.8-98.1) | 205 | 54.8 (49.1-60.6) | 26 | 6.9 (4.1-9.8) | 126 | 33.5 (27.9-39.1) | 14 | 3.8 (2.1-5.5) |
| 45-54 | 387 | 373 | 96.4 (94.5-98.2) | 235 | 60.6 (55.1-66.0) | 19 | 4.9 (2.8-7.0) | 148 | 38.2 (33.2-43.3) | 10 | 2.5 (0.9-4.1) |
| 55-65 | 364 | 345 | 94.8 (92.5-97.0) | 215 | 59.2 (53.1-65.3) | 21 | 5.7 (3.0-8.4) | 145 | 40.0 (33.1-46.9) | 11 | 2.9 (1.3-4.6) |

<sup>1</sup>-All estimates corrected for sampling design effect

A total of 1,043 participants had previous ZIKV infection (anti-ZIKV IgG-ELISA), yielding an overall weighted seroprevalence of 50.4% (95%CI: 47.2%-53.6%) (Table 2, Fig 3) and a sensitivity and specificity adjusted prevalence of ZIKV infection (anti-ZIKV NS-1 IgG and/or IgG3) of 38.6% (95%CI: 22.8%-54.2%) (Table 3). According to socioeconomic strata, the weighted Zika seroprevalence (IgG-ELISA) was statistically significantly lower in the high SES when compared to the intermediate and low strata, (Table 2, Fig 3). The weighted seroprevalence of recent ZIKV infection (anti-ZIKV IgG3) was 5.0% (95% CI: 3.9%-6.2%); with small variations from 4.0% to 6.9% in the different age groups (Table 2). The sensitivity and specificity adjusted seroprevalence of recent infection (anti ZIKV-IgG3) was 1.5% (95% CI: 0.1 % -3,7%) (Table 3).

**Table 3.**
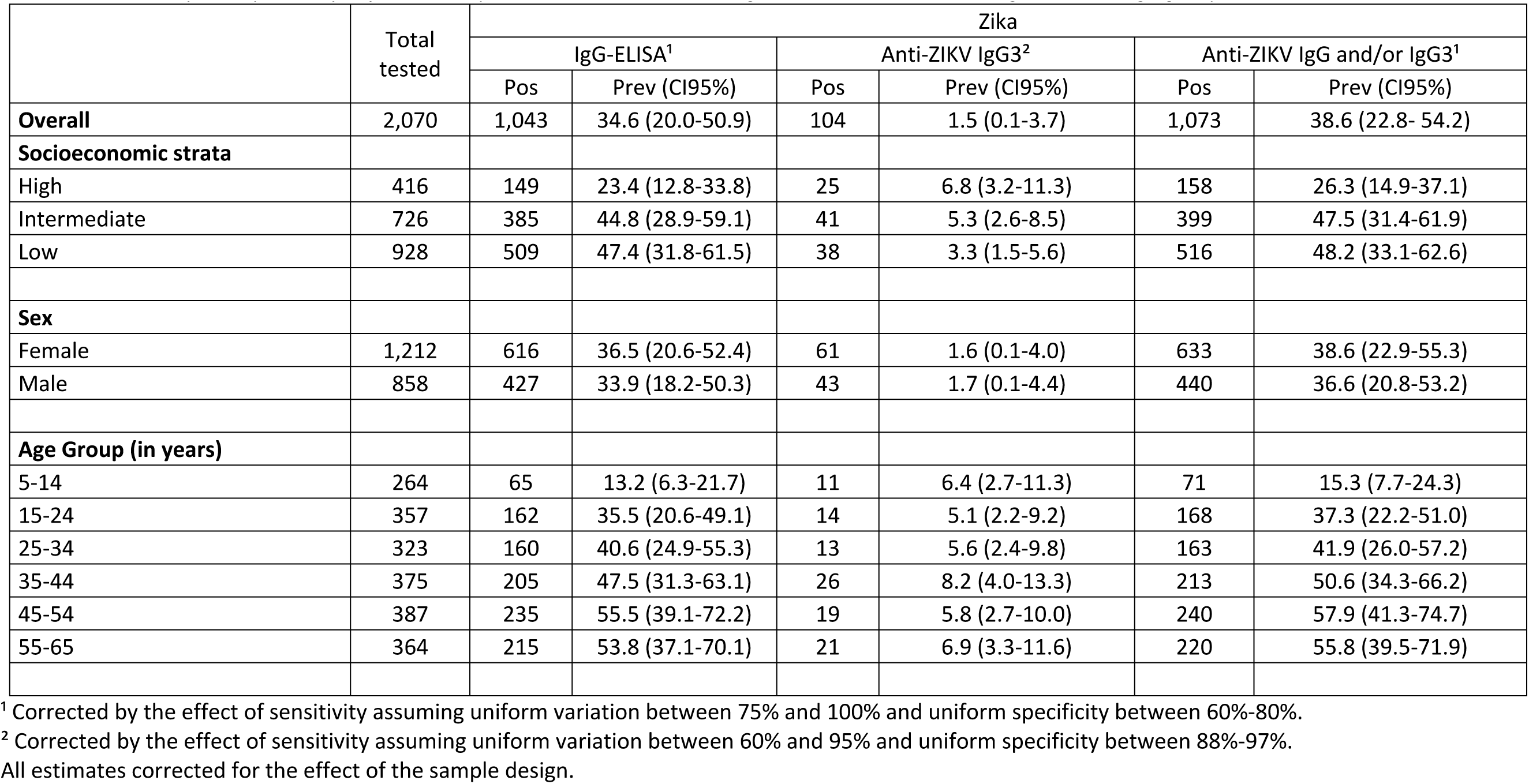
–Sensitivity and specificity adjusted seroprevalence of Zika according to socioeconomic strata, gender and age groups. Recife, Northeast of Brazil, 2018-2019.

In the female population of reproductive age, the seroprevalence of Zika corrected by the design effect was 52.8% (95%CI: 48.5%-57.1%). The seroprevalence of Zika (anti-ZIKV NS-1 IgG and/or IgG3) adjusted by the test accuracy was 44.8% (95%CI: 29.5%-59.2%).

The weighted seroprevalence for ZIKV among the children (<15 years old) was 24.6% (95%CI:18.4-30.9%); which is significantly lower than that for young adults (14-24 years) with 45.3% (95%CI:38.8-51.9%) (Table 2).

A total of 770 participants had markers of previous CHIKV infection (anti-CHIKV IgG and/or IgM), resulting in an overall weighted seroprevalence of 37.2% (95% CI: 34.0% - 40.4%) (data not shown on the tables). The prevalence of previous CHIKV infection (anti-CHIKV IgG) was significantly lower in the high SES when compared to the intermediate and low strata. The prevalence of recent CHIKV (anti-CHIKV IgM) infection was 3.5% (95%CI: 2.7%-4.2%), with no difference between strata (Table 2, Fig 3). Females and males had similar markers of past and recent infection for dengue, Zika and chikungunya. Unlike to what was observed with chikungunya, the seroprevalence of dengue and Zika increased significantly with age (Table 2 and 3)

### Force of infection of dengue, Zika and chikungunya

Fig 4 and Table 4 show the age-related force of DENV, ZIKV and CHIKV infections estimated through generalized linear models (GLM) according to SES and using the serostatus (recent and/or previous infection) as the outcome variable. The seroprevalence of dengue, Zika and chikungunya showed non-linear associations with age in the three SES. Differently from what was observed in the seroprevalence of dengue and Zika, there was no statistically significant association of chikungunya seroprevalence with age.

**Figure 4.**
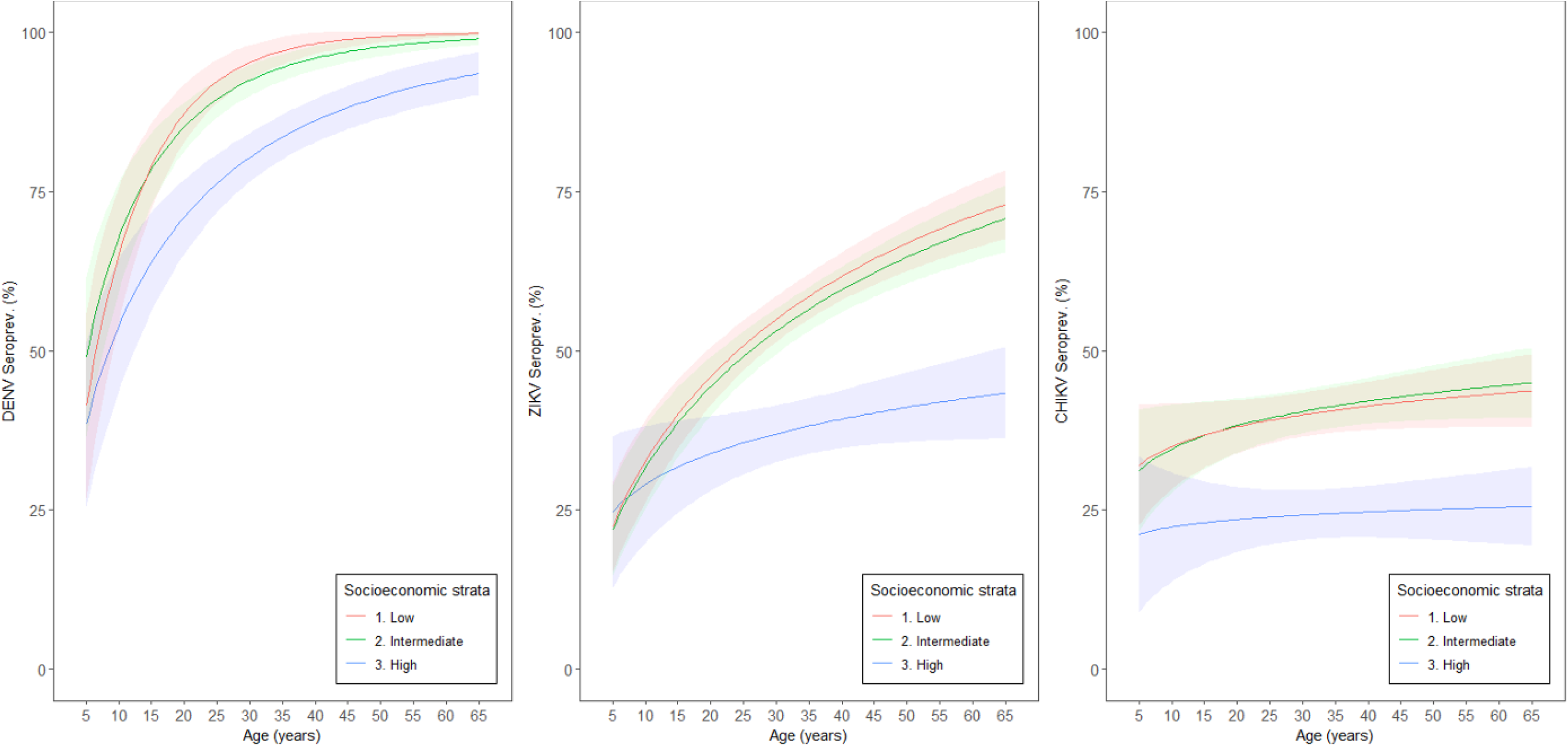
– Force of infection of Zika, Dengue and Chikungunya by according to socioeconomic strata. Recife, Brazil.2018-2019.

**Table 4.** Force of dengue, Zika and chikungunya infection (fitted by the effect of the sample design) through a generalized linear model (GLAM).

|  | High |  | Intermediate |  | Low |  |
| --- | --- | --- | --- | --- | --- | --- |
|  | Estimate | p-value | Estimate | p-value | Estimate | p-value |
| **Dengue** |  |  |  |  |  |  |
| Intercept | -1.8102 | <0.001 | -1.6031 | <0.001 | -2.1936 | <0.001 |
| Log(age) | 0.6753 | <0.001 | 0.7506 | <0.001 | 0.9736 | <0.001 |
| **Zika** |  |  |  |  |  |  |
| Intercept | -1.6993 | 0.004 | -2.4131 | <0.001 | -2.4040 | <0.001 |
| Log(age) | 0.2720 | 0.078 | 0.6274 | <0.001 | 0.6402 | <0.001 |
| **Chikungunya** |  |  |  |  |  |  |
| Intercept | -1.5682 | 0.003 | -1.2781 | <0.001 | -1.2011 | 0.003 |
| Log(age) | 0.0835 | 0.508 | 0.1828 | 0.079 | 0.1551 | 0.133 |

The seroprevalence of dengue, which had been circulating for 4 decades in the city, ranged from about 50% at 5 years of age and reached levels close to 100% in the low socioeconomic stratum - and these levels have been stable for more than 10 years. We observed that the seroprevalence curve in the high socioeconomic stratum or areas? reached a plateau at around 50 years of age, while in the intermediate and low SES, the plateau already occurred at around 30 years of age. The estimated force of infection was estimated to be 1.4 times higher in the low SES compared to the high SES.

Similarly, for ZIKV infection, the estimated force of infection was 2.4 times greater in the low socioeconomic strata when compared to the high strata. The seroprevalence of infection in the high SES, which was 30.2% at 5 years of age, reached a level of 44.6% at 65 years of age. In the intermediate and low SES, respectively, ZIKV seroprevalence, which was 33.9% and 32.5% at 5 years of age, reached levels of 73.0% and 76.8% at 65 years of age. This data suggests that high ZIKV herd immunity in adults living in the low SES might be the main factor contributing to the cessation of the ZIKV epidemic and not by the whole population average.

The seroprevalence of chikungunya was 21.6% at 5 years of age and increased to 26.8% at 65 years of age in the high SES. In the intermediate and low SES, the seroprevalence was around 35% at 5 years, and reached up to approximately 45% at 65 years in both strata (Fig 4)

Associated risk factors for ZIKV and CHIKV infection.

The supplementary Tables 5 to 10 (S4 supporting information) show the results of the crude and adjusted analysis of the association of individual and household characteristics with ZIKV and CHIKV infection in the distinct SES.

Regarding ZIKV infection, older age and living in a house were independent risk factors for infection in the three socioeconomic strata. A dose-response gradient of the risk of exposure to ZIKV with increasing age was observed in the intermediate and low SES. Living in a house represented a 2-fold increased risk of infection in the three SES. Presence of anti-dengue antibodies represented a 3-fold increased risk for ZIKV infection among participants from the high socioeconomic stratum. Low education was a risk factor for infection at the individual level in the high and intermediate strata, although with weak strength of association. Higher individual and head of household income were a protective factor against ZIKV infection in the intermediate SES (Table 7 – S4 supporting information).

Tables 8 to 10 (S4 supporting information) show the results of crude and adjusted analyzes of the association of household and individual factors with CHIKV infection. The risk of CHIKV infection was approximately three times higher among those living in a house compared to those individuals living in apartments in the high (adjusted OR 2.64; 95%CI: 1.34-5.17) and intermediate (adjusted OR 3.23; 95% CI: 1.80-5.50) SES. Most of the participants in the low SES lived in households (96.8%), and no CHIKV positive was detected among those who lived in an apartment in this area.

Considering the household-related characteristics associated with CHIKV infection, lack of access to the public sewage system and the lower level of schooling of the head of the household were risk factors for infection in the high SES. There was a negative association between the head of the family’s income and risk of CHIKV infection in the low stratum (Table 10 – S4 supporting information).

In the intermediate SES, the presence of serological marker of DENV infection represented a risk of CHIKV infection twice as high when compared to those without infection (adjusted OR 1.97; 95%CI: 1.28-3.02).

In the high SES, participants with fundamental/illiterate level of education had higher risk of CHIKV infection when compared to those with university level. The report of daily use of repellent was a protective factor in relation to those who did not use it (adjusted OR 0.22; 95%CI: 0.05-0.87). This use of repellent was not associated with infection in the other strata (Table 10 – S4 supplementary material).

## Discussion

This population-based survey estimated dengue, Zika, and chikungunya seroprevalence in a hyperendemic urban area of dengue and the epicenter of the Zika-related microcephaly epidemic in Brazil. The study, conducted after nearly three decades of DENV circulation and two years after the introduction of ZIKV and CHIKV, confirmed the intense transmission of arboviruses in this setting. After the first epidemic wave of ZIKV and CHIKV, about 50% and 30% of the population residing in the city, had exposure markers to these arboviruses, respectively, demonstrating the high vulnerability of this population to diseases transmitted by *Aedes aegypti*. The study also provided evidence of the persistence of (low-level?) co-circulation of ZIKV and CHIKV, measured by serological markers of recent infection (anti-ZIKV IgG3 and anti-CHIKV IgM), during an interepidemic period as reported case for both diseases, according to the official surveillance system in this setting [40].

Our study has some limitations inherent of serosurveys in large urban settings. First, children below five years of age were not eligible due to the difficulty of obtaining blood specimens during household visits. As expected in large urban areas, some dwellings could not be accessed and/or we experienced a rate of refusal in the high socio-economic stratum compared to the other strata. Nevertheless, the data presented are based on a population-based study using a probabilistic sampling design. The serologic tests of Zika were optimized and calibrated using a validated panel of well characterized dengue and Zika samples to correlate PRNT data with antibody binding data and determination of cut-off levels. We further adjusted the prevalence of Zika for a Bayesian model to account for the sensitivity and specificity of the Elisa test.

In a previous population-based survey in the city of Recife around 15 years ago (2006), we estimated that almost the entire population had DENV IgG antibodies for one or more DENV serotypes [12, 28]. A small hospital-based study in pregnant women found similar levels of almost 100% of DENV exposure in 2010 in this population [41]. This level of seroprevalence could be classified as one of the highest DENV seroprevalence rates worldwide; only comparable to other highly endemic countries in America, such as Trinidad, French Caribbean, Ecuador, and Suriname [42, 43].

In this population-based survey conducted in 2018-2019, the overall ZIKV seroprevalence (50.4%) suggests an intense virus exposure during the first wave of ZIKV epidemic in 2015-2016 in the municipality of Recife, Northeast Brazil. This prevalence may be considered high even when adjusting for the non-optimal accuracy of the serological test performed (38.6%). We highlight that the adjusted seroprevalence for women at reproductive age reached close to 45% of ZIKV infection in this population. This high level of ZIKV exposure may explain one of the highest prevalence of adverse outcome of pregnancy such as microcephalic cases reported during the first wave of epidemic in this setting [15]. The results of different studies may not be comparable due to the distinct laboratory tests applied and/or study designs. In our setting, a case-control study found 57.2% of seroprevalence of ZIKV virus by PRNT among pregnant women with non-adverse outcomes (control sample) during the peak of the epidemic [44]. Netto and al [45] reported high ZIKV seroprevalence (63%) using Elisa and PRNT tests among a convenience sample in Salvador, Bahia, estimating a reproduction number of 2.1 during the outbreak.

The levels of ZIKV seroprevalence found in our study are in line with results reported in the adult population in large urban center of Nicaragua after 2016 epidemic wave. Other international surveys post epidemic peak pointed out to higher seroprevalence levels such as the results reported after outbreaks in Yap Island [46], Pacific Region and French Polynesia, Micronesia [47]. Interestingly, we found at least 1.5% prevalence of recent marker of infection (IgG3) suggesting the persistence of ZIKV circulation in our setting after the 2015/2016 epidemic. This finding is corroborated by the report of the Pan American Health Organization of almost 30,000 cases in 2022. However, the incidence of ZIKV viremia using molecular tests was undetectable among blood donors in the same setting and period of our study [48].

The likely explanation may be the short duration of viremia in a self-selected healthy population as blood donors. However, our results support that ZIKV has been circulating at lower levels even after a large epidemic. ZIKV circulation after a large outbreak has been a matter of discussion in the recent literature [49] and deserves further population-based studies.

In our densely urbanized setting with high infestation of *Aedes aegypti*, the CHIKV displaced ZIKV outbreak (2015-2016) [5]. The current survey showed high prevalence of CHIKV infection (35.7%), with little variation among age groups as expected considering a recently introduced virus. This level of infection was not sufficient to avoid the occurrence of another CHIKV epidemic detected (2021) by the official surveillance, approximately one year after our survey [50]. In a meta-analysis of CHIKV seroprevalence studies conducted in Brazil, the estimated overall prevalence was 24% including only three studies [43].

Another interesting finding was 3.5% prevalence of recent CHIKV infection and the simultaneous co-circulation of CHIKV and ZIKV in our setting. Concurrently to this survey, we also described the co-circulation of ZIKV and CHIKV among pregnant women in a maternity-based study conducted in this same city [51]. In fact, the circulation of CHIKV during ZIKV (2015-2017) outbreak was previously documented among pregnant women with rash notified by the official surveillance system in our setting [52].

The introduction of CHIKV in the Northeast of Brazil is quite recent compared to other regions such as Southeast Asia where this virus has been circulating for several decades [53]. Our study showed evidence that one third of the population, i.e., around 544 thousand people living in the city had been exposed to CHIKV since the first epidemic wave (2016) [54]. This figure is 45-fold higher than the 11,984 accumulated reported CHIKV suspected cases reported by the surveillance system since 2015[50], until the end of the current survey (Feb. 2019). This estimated ratio between CHIKV infection and notified cases suggests substantial case underreporting and/or high frequency of subclinical/inapparent infections.

We did not find statistical differences between the sexes regarding the seroprevalence of the three arboviruses surveyed. This result is in accordance with previous population surveys conducted in this setting [12], in other Brazilian states [10, 55] and other countries in the region [56]. Conversely, population-based studies conducted in the Americas reported higher seroprevalence of ZIKV and CHIKV in males compared to females [57, 58]. This difference can be explained by local characteristics of the population or by methodological differences across the studies.

Interestingly, our analysis showed different age-related infection curves for dengue, Zika or chikungunya. At 5 years of age, approximately 50% of the children had markers of previous DENV infection and that more than 90% of the adult population had been infected. These findings are in consonance with the intense circulation of the virus in this population for four decades. A similar age-related pattern for dengue infection was also documented by our research team in this setting in 2006 [12, 29]. In addition, we observed a steady increase in the force of ZIKV infection from 25% of prevalence at the age of 5 years to 70% at the age of 65, confirmed by the results of the regression model (Table 4 and Supplementary Material S1). This finding was consistent with the results of the multiple regression analysis, which, unlike that observed for CHIKV infection, showed a higher risk of infection in the population aged 15 years and older when compared to the population below this age range. Although this finding seems unexpected for a recent introduced virus, it is in line with results from Nicaragua [57] and Puerto Rico [58]. The significant increase in the seroprevalence between children and young adults may suggest that anti-dengue antibodies or sexual transmission may have influenced the transmission of Zika. Seroprevalence levels around 60-70% which suggest herd immunity are only found in (older) adults of low SES, suggesting this group as the major driver for population immunity. In addition, higher levels of anti-dengue antibodies that is found in adults after multiple dengue exposures can provide partial protection against ZIKV also contributing to the overall herd immunity. Other studies have reported a significant role of sexual transmission of Zika [59–61], which could also contribute to the higher seroprevalence in (older) adults. Another population survey conducted in French Guiana did not show evidence of increased seroprevalence with age [56], and it still controversial if sexual transmission plays a significant role for ZIKV epidemiology[61, 62]. In contrast, the age-related curve of infection for CHIKV was estimated almost as a straight line that is compatible with the recent introduction virus in this region.

The weighted seroprevalence of dengue, zika and chikungunya was higher in areas classified as low and intermediate socioeconomic levels when compared to the high socioeconomic strata which have greater coverage of sanitation, regular water supply and where most of the population live in apartment buildings. In general, our data also reinforce the role of unplanned urbanization and poverty as one of the factors that influence the incidence and expansion of arboviruses [63, 64].

The analysis of the association between individual and household characteristics and dengue, ZIKV and CHIV infection showed different patterns. ZIKV and CHIKV infections were associated with lower educational levels as an indicator of health inequities and an independent risk factor for infection in almost all socioeconomic strata. In addition, living in a household instead of an apartment yielded a three-fold increased risk of exposure to CHIKV or to ZIKV infection. We also found that living in a house compared to high rise flat was a risk factor for DENV infection in the previous survey (2005/2006) in the city of Recife [12]. These findings are in line with results from other surveys in localities in southeastern Brazil [65, 66] and in the city of Singapore in Asia [67, 68]. In our setting, the greater risk of exposure to these arbovirus infections among residents of houses (one-story dwellings) can be explained by the fact that, unlike the areas of the high stratum, most of the households located in the intermediate and low socioeconomic strata consist of this type of residence (more than 80%). These densely populated areas, with less sanitation coverage and irregular water supply, provide the most favorable environmental conditions for the proliferation of *Aedes* breeding sites [63]. Another possible explanation would be the greater proximity to Aedes breeding sites in a household compared to high rise flats located above the ground floor.

In conclusion, our household-survey highlights the high vulnerability of these urban population to *Aedes*-borne arbovirus infections in the Northeast region of Brazil. Also, our results provide evidence of the persistence of the co-circulation of ZIKV and CHIKV in this highly urbanized setting, three years after the peak of ZIKV epidemic.

Considering the large proportion of susceptible population for ZIKV and CHIKV infection, it should be an alert for future outbreaks. There is an urgent need of new approaches to *Aedes* surveillance and control, besides the development and efficacy trials of vaccines against these arboviruses. The planning and implementation of intersectoral and integrated interventions for the control of diseases transmitted by *Aedes* in poor urban setting should be crucial to reduce the burden of arbovirus disease and mortality in endemic countries.

## Data Availability

All relevant data are within the manuscript and its Supporting Information files. Additional information will be provided upon request

## Acknowledgments

We thank Andre Sá de Oliveira for generating the figure 1. We also thank Dylan Tuttle and Dr. Priscila M. Castanha for reviewing the text and making figures 2 and 3.

## Conflicts of interest

We declare no conflict of interest.

## Supporting information

S1 Laboratory procedures

S2 Table. Exposure variable list

S3 Packages for Prevalence Data Analysis using R Programme

S4 Table 5 to 10-Univariate and adjusted analysis of the association between household and individual characteristics with ZIKV and CHIKV infection. Recife, Brazil, 2018-2019.

## Author contributions

Conceptualization: CB, WVS, MFPMA, CFCAM, TJ, ETAM, CMTM

Data curation: CB, RDL, WVS, CFL, CAM, IFTV

Formal analysis: CB, RDL, WVS, CFL, CMTM, IFTV

Funding acquisition: CB, ETAM, TJ, IFTV

Investigation: CB, CAM, CNLM, CAAB, CFCAM, ETAM, IFTV, JFD

Methodology: CB, WVS, CFL, CFCAM, ETAM, CMTM, IFTV

Project administration: CB, RDL, TJ, JFD, IFTV

Supervision: CB, RDL, CAM, CNLM, IFTV

Validation: IFTV

Writing – Original Draft Preparation: CB, WVS, CFL, MFPMA, CAM, RDL, TJ, CMTM, IFTV, JFD

Writing – Review & Editing: CB, MFPMA, CNLM, CAAB, CFCAM, RDL, TJ, ETAM, CMTM, IFTV.

## Notes

### Competing Interest Statement

The authors have declared no competing interest.

### Funding Statement

Funding: European Commission (ZIKA Alliance H2020 734548) German Centre for Infection Research (DZIF), Heidelberg Site, Germany Pan American Health Organization, World Health Organization/Brazilian Ministry of Health (SCON2018-00276) National Council for Scientific and Technological Development, Brazil (CNPq scholarship – CB (303953/2018-7), WVS (308000/2021-8), MFPMA (CNPq 302696/2021-0)

### Author Declarations

The research project was reviewed and approved by the Research Ethics Committee of the Aggeu Magalhães Institute (Fiocruz, Pernambuco) (CAEE: 79605717.9.0000.5190, report number 2.734.481).

